# Nonspecific blood tests as proxies for COVID-19 hospitalization: are there plausible associations after excluding noisy predictors?

**DOI:** 10.1101/2020.10.12.20211557

**Authors:** G. Ishikawa, G. Argenti, C. B. Fadel

**Affiliations:** Professor and researcher at Universidade Tecnologica Federal do Parana (UTFPR) in Ponta Grossa, Brazil; Researcher enrolled in the postgraduate programme in health sciences at Universidade Estadual de Ponta Grossa (UEPG) in Ponta Grossa, Brazil; Professor and researcher at Universidade Estadual de Ponta Grossa (UEPG) in Ponta Grossa, Brazil

**Keywords:** COVID-19, Prediction, Hospitalization, Laboratory tests, Creatinine, Eosinophils, Monocytes, Neutrophils, C-protein reactive

## Abstract

This study applied causal criteria in directed acyclic graphs for handling covariates in associations for prognosis of severe COVID-19 (Corona virus disease 19) cases. To identify nonspecific blood tests and risk factors as predictors of hospitalization due to COVID-19, one has to exclude noisy predictors by comparing the concordance statistics (AUC) for positive and negative cases of SARS-CoV-2 (acute respiratory syndrome coronavirus 2). Predictors with significant AUC at negative stratum should be either controlled for their confounders or eliminated (when confounders are unavailable). Models were classified according to the difference of AUC between strata. The framework was applied to an open database with 5644 patients from Hospital Israelita Albert Einstein in Brazil with SARS-CoV-2 RT-PCR (Reverse Transcription – Polymerase Chain Reaction) exam. C-reactive Protein (CRP) was a noisy predictor: hospitalization could have happen due to causes other than COVID-19 even when SARS-CoV-2 RT-PCR is positive and CRP is reactive, as most cases are asymptomatic to mild. Candidates of characteristic response from moderate to severe inflammation of COVID-19 were: combinations of eosinophils, monocytes and neutrophils, with age as risk factor; and creatinine, as risk factor, sharpens the odds ratio of the model with monocytes, neutrophils, and age.

## INTRODUCTION

COVID-19 (Corona virus disease 19) caused by SARS-CoV-2 (acute respiratory syndrome coronavirus 2) stands out for its high rate of hospitalization and long hospital stay and in intensive care units (ICU). COVID-19 disease severity can be mild, moderate, severe, and critical [1]. While 81% of those infected with COVID-19 have mild or moderate symptoms, World Health Organization estimates that 14% of those infected with COVID-19 are severe and require hospitalization and oxygen support, and 5% are critical and admitted to intensive care units [1]. Reported median hospital length of stay (LoS) was from 4 to 21 days (outside China) and ICU LoS was from 4 to 19 days [2].

The severity of COVID-19 states is associated with many risk factors. Early reports suggest advanced age, morbidities, multi-morbidities, and immunosuppression [3,4]. The enlarging list includes cardiac, chronic lung, cerebrovascular, chronic kidney, and liver diseases, cancer, diabetes, obesity, hypertension, dyspnea, fatigue, and anorexia [1,5,6].

Early identification of severe cases allows for optimizing emergency care support [1] and improving patient outcomes [7]. However, patients who do not yet meet supportive care criteria may fail to receive the necessary care, when there is rapid deterioration or inability to promptly go to a hospital. In the transition from moderate to severe cases there can be avoidable delays in life support interventions with non-optimized treatments.

Together with high hospitalization rates [1] and lengthy stay [2], the superposition of COVID-19 waves and sustained transmission [8] are causing prolonged depletions of health care resources in many countries. Prognosis tools may play a role in planning and in improving the access to supportive treatments by allowing timely allocation of scarce resources to better cope with COVID-19. Indeed, there is widespread interest in predictive models of COVID-19 outcomes [7,9], but a review of 50 prognostic models concluded that they are at high risk of bias [9]. As they focus on statistical findings, our concern is with lack of minimum causal criteria to identify associations that are effectively related to COVID-19.

In this context, a path to optimized supportive treatments is more reliable assessments of the transition from moderate to severe cases of COVID-19 inflammation. We choose nonspecific blood tests as they are widely available, and hospitalization decision as a proxy to characterize the transition from moderate to severe cases (when not constrained by inpatients availability). After formalizing an analytical framework with causal reasoning, the goal is to identify candidate sets of blood tests associated with hospitalization (with risk factors), excluding noisy predictors that are not related to COVID-19 inflammation.

## METHODS

Whereas causal effects are clearly predictive, prediction studies usually refer to noncausal analysis that uses observational data to make predictions beyond the observed ones and confounding bias is generally considered a nonissue [10]. But when one needs more reliable predictions, confounding bias and causality should be accounted for in associations. This study applies analytical tools from the causal effect estimation of directed acyclic graph theory [11] to investigate associations considering covariates.

The strength of the association depends on the specificity and sensitivity of the inflammation pattern, as a kind of distinctive signature of the disease. A low association can also occur and means that the pattern with that set of variables allows weak inferences. If a substantial association due to COVID-19 is identified and it is also stable and representative of the target population, then these blood tests may be useful as proxies in surveillance protocols and screening interventions.

### Theoretical framework

The theory of directed acyclic graph (DAG) provides graphical notation and a non-parametric probabilistic terminology to describe and evaluate causal relationships [11]. The use of DAGs in epidemiology is emergent [12] and it is especially helpful with multiple potential confounders [12,13] that may introduce systematic bias [10,14]. In DAGs, confounding associations between two variables may come from unblocked backdoor paths [13] that can be graphically identified because they share parent nodes. With a formal definition of backdoor path, for instance, DAG provides a general explanation of the Simpson’s paradox [15], where a phenomenon appears to reverse the sign of the estimated association in disaggregated subsets in comparison to the whole population. As a framework, DAG supplies analytical tools to evaluate which adjustment is mandatory (to predict a noncausal sign reverse) and which covariate should be omitted (to estimate the causal effect), thereby enforcing the elicitation of qualitative causal assumptions [11,12,14].

A hypothetical DAG model with latent variable was conceived to evaluate the influence of various types of covariates on the focal association. Initially we drew the main causal path from exposure to outcome. The DAG in Figure 1 starts from the infection by SARS-CoV-2 (exposure E) that, in some cases, leads to “Moderate to Severe Inflammation due to COVID-19” (MSIC, hypothetical latent variable [E→MSIC]), and that inflammation causes two outcomes (mutual dependent relationship [H←MSIC→B]): (H) hospitalization decision; and (B={B_1_,…,B_k_}) blood tests measured at hospital admission. The blood tests are selected according to their strength with hospitalization. The focal outcomes under investigation are hospitalization (H) and blood tests (B).

**Figure 1.**
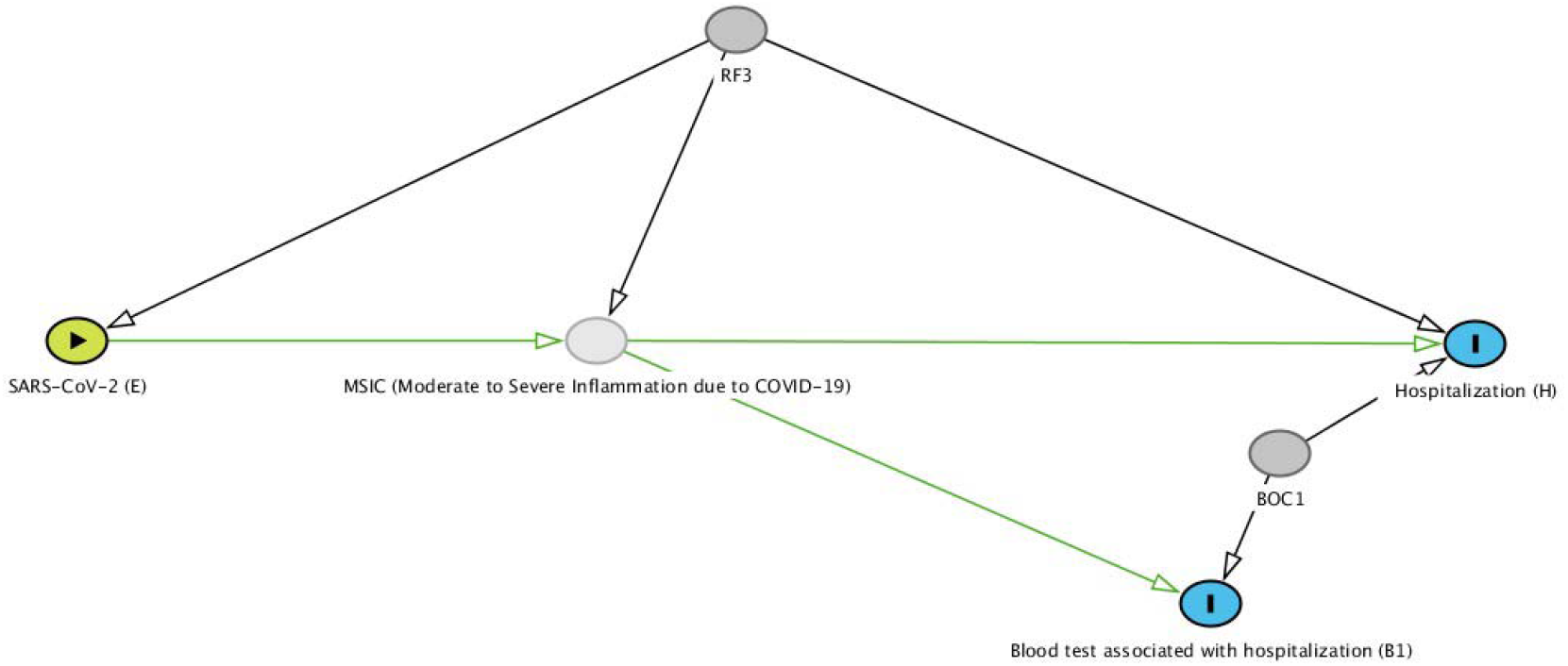
Initial hypothetical directed acyclic diagram with the main causal path of a moderate to severe COVID-19 inflammation (MSIC), one risk factor (RF3) and one confounder (BOC1) of the focal outcomes (H and B1). MSIC is a latent variable (unmeasured); Outcomes are H: hospitalization (H={regular ward, semi-intensive care, intensive care unit}); and B: blood test (B={B_1_})

Considering the initial DAG plausible, we hypothesized candidate covariates that are parents of the variables and may open back-door paths, Figure 1 shows one risk factor (RF3) and one confounder (BOC1). Figure 2 is an enhancement of the initial DAG with potential risk factors, confounders of the focal association, and other covariates. Risk factors contribute directly to the development of COVID-19 inflammation (RF={RF_1_,…,RF_L_}, mutual causation relationships [RF_i_→MSIC←RF_j_]) and they can also affect other variables. Figure 2 also distinguishes the covariates in terms of their confounding potential on the association between H and B. Covariates that affect both focal outcomes are identified as Both-Outcomes-Confounders (BOC={BOC_1_,…,BOC_m_}), as they are correlated to the focal outcomes but not to COVID-19, and when affect one outcome as Single-Outcome-Covariate (SOC={SOC_1_,…,SOC_n_}). These covariates are not exhaustive but to generate causal graph criteria for handling confounding factors.

**Figure 2.**
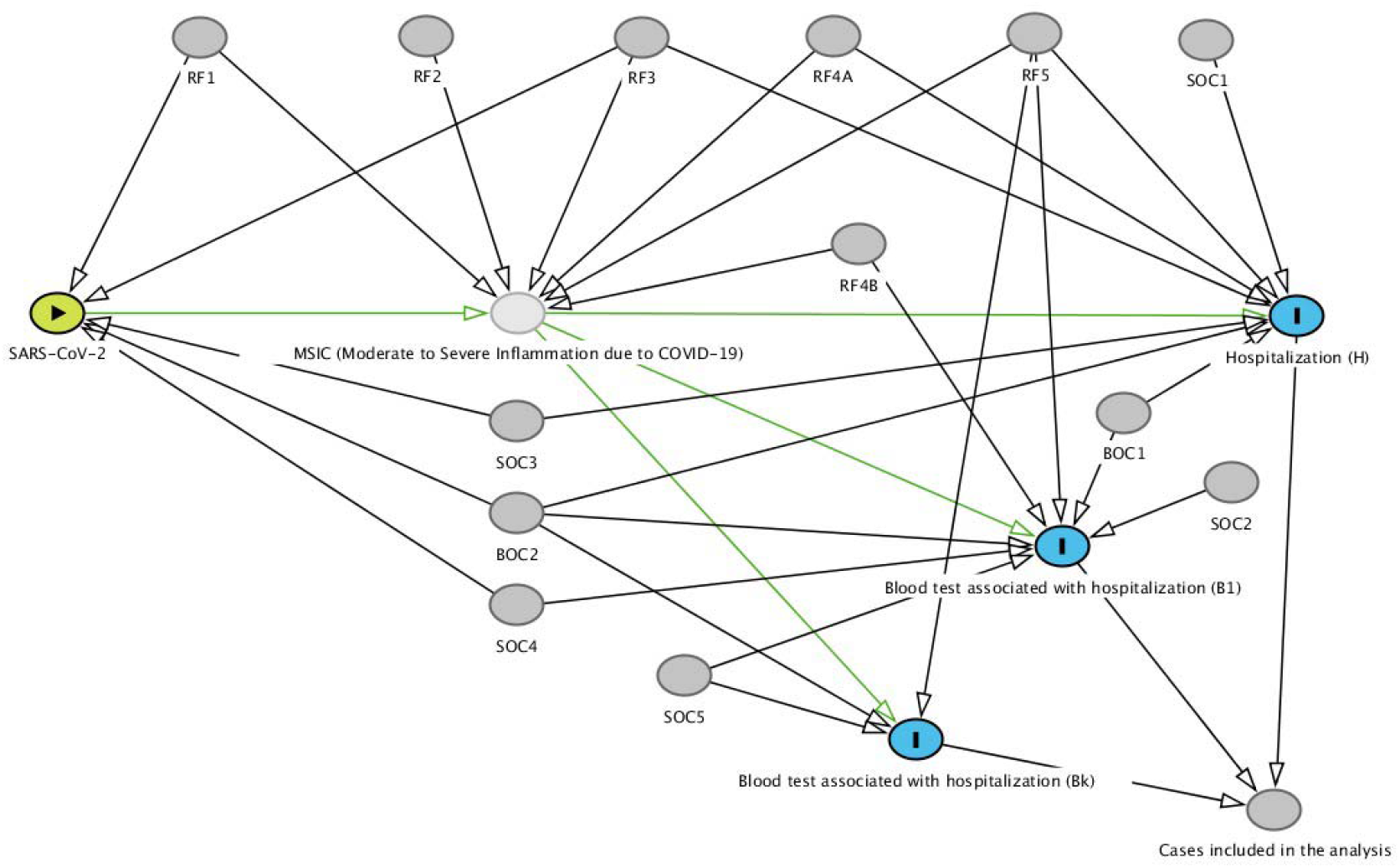
Hypothetical directed acyclic diagram of a COVID-19 inflammation causal path with risk factors, confounders and other covariates Exposure = SARS-CoV-2 (E) (acute respiratory syndrome coronavirus 2); Outcomes are H: hospitalization (H={regular ward, semi-intensive care, intensive care unit}), and B: blood tests (B={B_1_,…,B_1_}); Covariates are RF: risk factor (RF={RF_1_,…,RF_4A_, RF_4B_,RF_5_}), SOC: single outcome covariate (SOC={SOC_1_,…,SOC_5_}), and BOC: both outcomes confounder (BOC={BOC_1_,BOC_2_})

Causal relationships in DAGs are defined with the *do*(.) operator that performs a theoretical intervention by holding constant the value of a chosen variable [11,16]. The association caused by COVID-19 inflammation can be understood as a comparison of the conditional probabilities of hospitalization (H) given a set of blood tests (B) under intervention to SARS-CoV-2 infection (*do*(SARS-CoV-2)=1) and intervention without infection (*do*(SARS-CoV-2)=0):

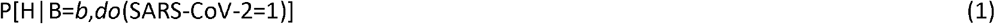

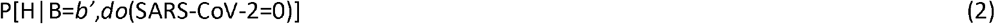

Where P[H|B=*b,do*(SARS-CoV-2=1)] represents the population distribution of H (hospitalization) given a set of blood tests equal to b, if everyone in the population had been infected with SARS-CoV-2. And P[H|B=*b’,do*(SARS-CoV-2=0)] if everyone in the population had not been infected. Of interest is the comparison of these distributional probabilities for each intervention.

The interventions with *do*(.) generate two modified DAGs:

- The *do*(SARS-CoV-2=0) eliminates all arrows directed towards SARS-CoV-2 and to MSIC (Figure 3). Ignoring the floating covariates, there are single arrow covariates pointing to hospitalization (RF3, RF4A, SOC1, SOC3) and to blood tests (RF4B, SOC2, SOC4) and fork covariates pointing to both outcomes (BOC1, BOC2, RF5).

**Figure 3.**
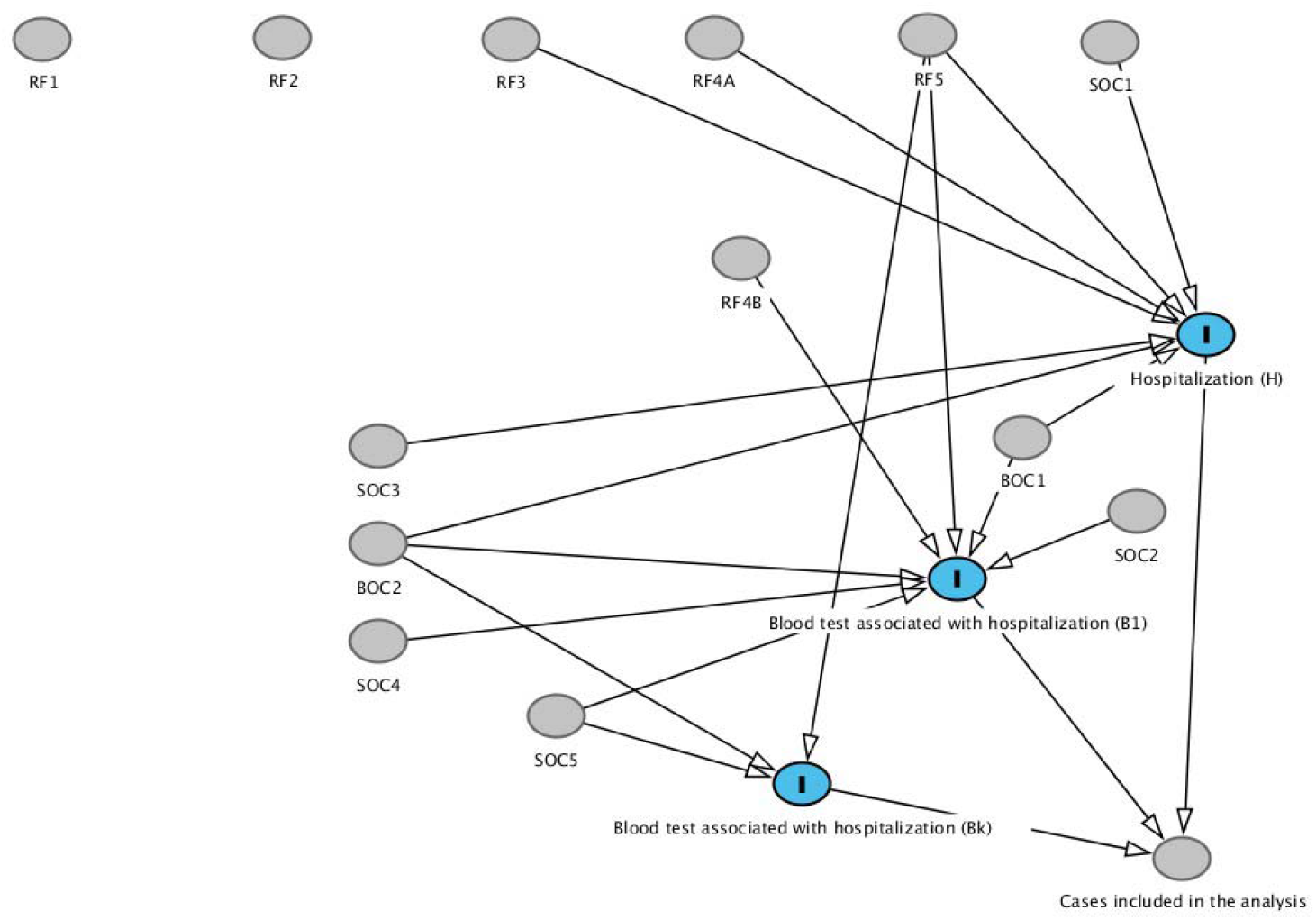
Modified directed acyclic diagram with intervention at no exposure (*do*(SARS-CoV-2=0)) to evaluate the influence of covariates on the focal outcomes (H and B) Exposure = SARS-CoV-2 (E) (acute respiratory syndrome coronavirus 2); Outcomes are H: hospitalization (H={regular ward, semi-intensive care, intensive care unit}), and B: blood tests (B={B_1_,…,B_K_}); Covariates are RF: risk factor (RF={RF_1_,…,RF_4A_, RF_4B_,RF_5_}), SOC: single outcome covariate (SOC={SOC_1_,…,SOC_5_}), and BOC: both outcomes confounder (BOC={BOC_1_,BOC_2_})
- Similarly, the modified graph of *do*(SARS-CoV-2=1) is equal to the former by adding single arrows from RF1 and RF2 to MSIC; and converting RF3, RF4A, RF4B, and RF5 to fork types with arrows directed to MSIC.

As most covariates are either unmeasured or unknown, the effect of their absence can be evaluated following the d-separation concept [11]. This concept attempts to separate (make independent) two focal sets of variables by blocking the causal ancestors (or back-door paths) and by avoiding statistical control for mutual causal descendants [11]. Differently, to preserve the association between descendants of MSIC (Figure 2), the focal outcomes (H and B) must remain d-connected (dependent on each other only through MSIC) and their relations with other covariates (that may introduce systematic bias) should be d-separated (conditionally independent). Figure 3, at the negative stratum, shows the confounders that may introduce systematic bias into both outcomes: BOC1, BOC2, RF5. The influence of these confounders on the focal association can be estimated with the modified model at the negative strata. A strong association of the outcomes without infection can be due to these confounders and suggest efforts to measure and control for them (as they have to be d-separated). Another pragmatic possibility is to exclude the noisy exams affected by these confounders. The other covariates are single arrows or they affect only one outcome (H or B) – their absence should not be critical because they are likely to be discarded due to poor discriminative performance.

### Model assessment with naïve estimation

A naïve estimation of equations (1) and (2) is to assume that they are equal to their conditional probabilities available in a given dataset at each stratum. The cost of this simplification is that the analysis is no longer causal (in a counterfactual sense, because we are not contrasting the whole population infected and the whole population not infected [10,11,16]) and the estimation becomes an association between two disjoint sets that each represents separate parts of the target population.

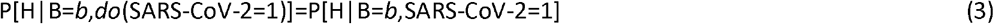

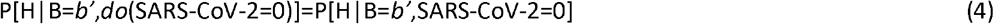

As Hospitalization is a dichotomous variable, this conditional probability, P[H|B=*b*,SARS-CoV-2=1], can be computed through a logistic regression of Hospitalization (dependent variable) given a set of blood tests at SARS-CoV-2=1. From the modified graph with intervention, P[H|B=*b’*,SARS-CoV-2=0] is calculated with the same model parameters but applied to cases at the negative stratum. It is implicit that there is the conditioning by a proper set of covariates for each model.

The concordance statistic of a logistic regression model is a measure of its predictive accuracy and is calculated as the Area Under of the receiver operating characteristic Curve (AUC) [10,17]. A way to compare the discriminative ability of (3) and (4) is to subtract the AUC values at each stratum. A difference of 0.0 means no specific association with COVID-19 (i.e., equivalent responses for both strata) and 0,5 means perfect focal association of the outcomes and perfect differentiation among strata (i.e., perfect response at the positive stratum and random response at the negative).

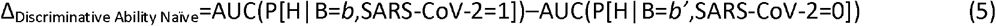

The comparison of the models with AUC values at the negative stratum of SARS-CoV-2 is a necessary improvement in the assessment of prognostic models. This is similar to the null values concept in measures of associations of two groups with two outcomes [10], but generalized for continuous multivariable prognostic models.

### Model selection criteria

The above framework guided our approach to identify sets of blood tests associated with the hospitalization due to COVID-19 together with:

- Acceptable overall statistical properties of each model at the positive stratum of SARS-CoV-2, without and with bootstrap procedure;
- Consistency of the blood test coefficients across models with one variable and with multiple variables: considering causal effects, coefficients should not change signal when properly conditioned across models [15]; and
- Elimination of models with high AUC at the negative stratum of SARS-CoV-2 and classification of the sets of blood tests by the difference of AUC between strata.

### Source dataset

We identified one public observational database in which, at least partially, we could apply the framework and generate candidate prognostic models. Hospital Israelita Albert Einstein (HIAE), Sao Paulo/Brazil, made public a database (HIAE_dataset)[18] in the *kaggle* platform of 5644 patients screened with SARS-CoV-2 RT-PCR (Reverse Transcription–Polymerase Chain Reaction) exam and a few collected additional laboratory tests during a visit to this hospital from February to March, 2020. All blood tests were standardized to have mean of zero and unitary standard deviation. As this research is based on public and anonymized dataset, it was not revised by any institutional board. The logistic regression models were evaluated with IBM SPSS version 22.0 and the causal map with DAGitty.net version 3.0.

## RESULTS

Of the 5644 patients, 558 presented positive results for SARS-CoV-2 RT-PCR. Of the 170 patients hospitalized (in regular ward, semi-intensive unit or intensive care unit), 52 were positive (9,3% rate of hospitalization due to COVID-19). Patient age quantile, from 0 to 19, with sample mean of 9,32, was the only demographic variable available. Age was not conditionally independent with SARS-CoV-2 RT-PCR exam. Only 0,9% were positive in the age quantile 0, 1, and 2 (8 positive cases in 883 exams) while the incidence (not weighted) in the age quantile from 3 to 19 was 11,7% ± 2,6%.

In the first round, 15 blood tests were discarded because of poor performance of the univariate model when SARS-CoV-2=1 (Table 1). The remaining blood tests were creatinine, C-Reactive Protein (CRP), eosinophils, lymphocytes, monocytes, and neutrophils (Table 1). Only creatinine was not related with the immune system directly and was evaluated as a risk factor. Of the 5644 patients, 602 patients presented values of eosinophils, 602 lymphocytes, 601 monocytes, 513 neutrophils, 506 CRP, and 424 creatinine. Regarding missing cases, all observations with the required data were included (available-case analysis).

**Table 1.** Univariate logistic regression models with blood tests for predicting hospitalization

|  | At SARS-CoV-2=1 |  |  |  |  |  | At SARS-CoV-2=0 |  |  |  |  |  |
| --- | --- | --- | --- | --- | --- | --- | --- | --- | --- | --- | --- | --- |
|  | N | B | p | OR | OR 95% C.I. |  | N | B | p | OR | OR 95% C.I. |  |
|  |  |  |  |  | Lower | Upper |  |  |  |  | Lower | Upper |
| zBasophils | 83 | -,374 | ,229 | ,688 |  |  | 519 | -,375 | ,010 | ,687 |  |  |
| zHematocrit | 83 | -,123 | ,658 | ,884 |  |  | 520 | -,976 | ,000 | ,377 |  |  |
| zHemoglobin | 83 | -,073 | ,785 | ,930 |  |  | 520 | -1,009 | ,000 | ,365 |  |  |
| zLeukocytes | 83 | ,617 | ,167 | 1,854 |  |  | 519 | ,658 | ,000 | 1,931 |  |  |
| zMCH | 83 | -,253 | ,280 | ,776 |  |  | 519 | -,289 | ,011 | ,749 |  |  |
| zMCHC | 83 | ,118 | ,629 | 1,126 |  |  | 519 | -,259 | ,023 | ,772 |  |  |
| zMCV | 83 | -,331 | ,176 | ,718 |  |  | 519 | -,196 | ,094 | ,822 |  |  |
| zMPV | 81 | -,465 | ,079 | ,628 |  |  | 518 | -,229 | ,062 | ,795 |  |  |
| zPlatelets | 83 | -,272 | ,433 | ,762 |  |  | 519 | ,101 | ,363 | 1,107 |  |  |
| zPotassium | 58 | -,482 | ,145 | ,618 |  |  | 313 | ,161 | ,210 | 1,174 |  |  |
| zRed_blood_cells | 83 | ,087 | ,707 | 1,091 |  |  | 519 | -,791 | ,000 | ,453 |  |  |
| zRDW | 83 | ,140 | ,560 | 1,150 |  |  | 519 | ,648 | ,000 | 1,912 |  |  |
| zSerum_glucose | 33 | -,172 | ,734 | ,842 |  |  | 175 | ,713 | ,001 | 2,041 |  |  |
| zSodium | 58 | -,530 | ,097 | ,589 |  |  | 312 | -,232 | ,077 | ,793 |  |  |
| zUrea | 59 | ,468 | ,275 | 1,597 |  |  | 338 | ,403 | ,004 | 1,496 |  |  |
| Age_quantile * | 558 | 0,199 | ,000 | 1,220 | 1,137 | 1,310 | 5086 | -0,03 | ,044 | 0,968 | 0,938 | 0,999 |
| zCreatinine ** | 62 | 1,002 | ,019 | 2,723 | 1,177 | 6,301 | 362 | -,116 | ,367 | ,891 | ,693 | 1,145 |
| zCRP ** | 70 | 1,857 | ,004 | 6,406 | 1,805 | 22,73 | 436 | 1,012 | ,000 | 2,751 | 2,015 | 3,756 |
| zEosinophils ** | 83 | -2,768 | ,001 | ,063 | ,012 | ,332 | 519 | -,312 | ,036 | ,732 | ,547 | ,980 |
| zLymphocytes ** | 83 | -,794 | ,006 | ,452 | ,256 | ,796 | 519 | -,537 | ,000 | ,584 | ,451 | ,758 |
| zMonocytes ** | 83 | -,629 | ,006 | ,533 | ,339 | ,838 | 518 | -,321 | ,021 | ,726 | ,552 | ,953 |
| zNeutrophils ** | 75 | 1,412 | ,000 | 4,104 | 1,957 | 8,605 | 438 | ,509 | ,001 | 1,663 | 1,244 | 2,224 |

SARS-CoV-2 (acute respiratory syndrome coronavirus 2): result of the exam for SARS-CoV-2 RT-PCR (0=negative; 1=positive) (reverse transcription – polymerase chain reaction)
N: Cases included in the analysis; B: coefficient of the univariate logistic regression; p: coefficient significance; OR: odds ratio (exp(B)); CI: confidence interval
MCH: Mean corpuscular hemoglobin; MCHC: Mean corpuscular hemoglobin concentration; MCV: Mean corpuscular volume; MPV: Mean platelet volume; RDW: Red blood cell distribution width
zName: means that the variable was converted and made available in a standardized format (mean=0; standard deviation=1)
\* Age was converted in quantiles in the range of 0 to 19, mean value is 9,32.
\*\* Blood tests selected for screening as potential predictors of COVID-19 inflammation

CRP is a biomarker of various types of inflammation [19,20]. At SARS-CoV-2=1, the model with CRP and age had good discriminative ability with AUC of, 872. But at SARS-CoV-2=0, AUC=,680 was also substantial and the difference of the discriminative ability Δ=,192 was moderate (candidate models should present higher differences); the corresponding ROC curve in Figure 4 show overlapping curves up to sensitivity of 0,5 to 0,6. Models with CRP demonstrated sensitivity to resampling within the dataset [17], the coefficient significance moved from, 005 to, 144. Similar effects were found in models that include CRP with other blood tests and sensitivity to bootstrapping was reduced by dichotomizing CRP (reactive/not-reactive). Models with CRP_reactive, neutrophils, and age generated AUC of, 901 and, 730 in the positive and negative strata (Δ=0,171), and CRP_reactive, monocytes, neutrophils, and age generated AUC of, 921 and, 706, respectively (Δ=,215). CRP is a predictor of hospitalization in general, but high levels of AUC at the negative stratum mean that CRP is a response with significant bias due to other causes than COVID-19. Differently from other prognostic studies [21,22,23,24,25,26], CRP was excluded as candidate.

**Figure 4.**
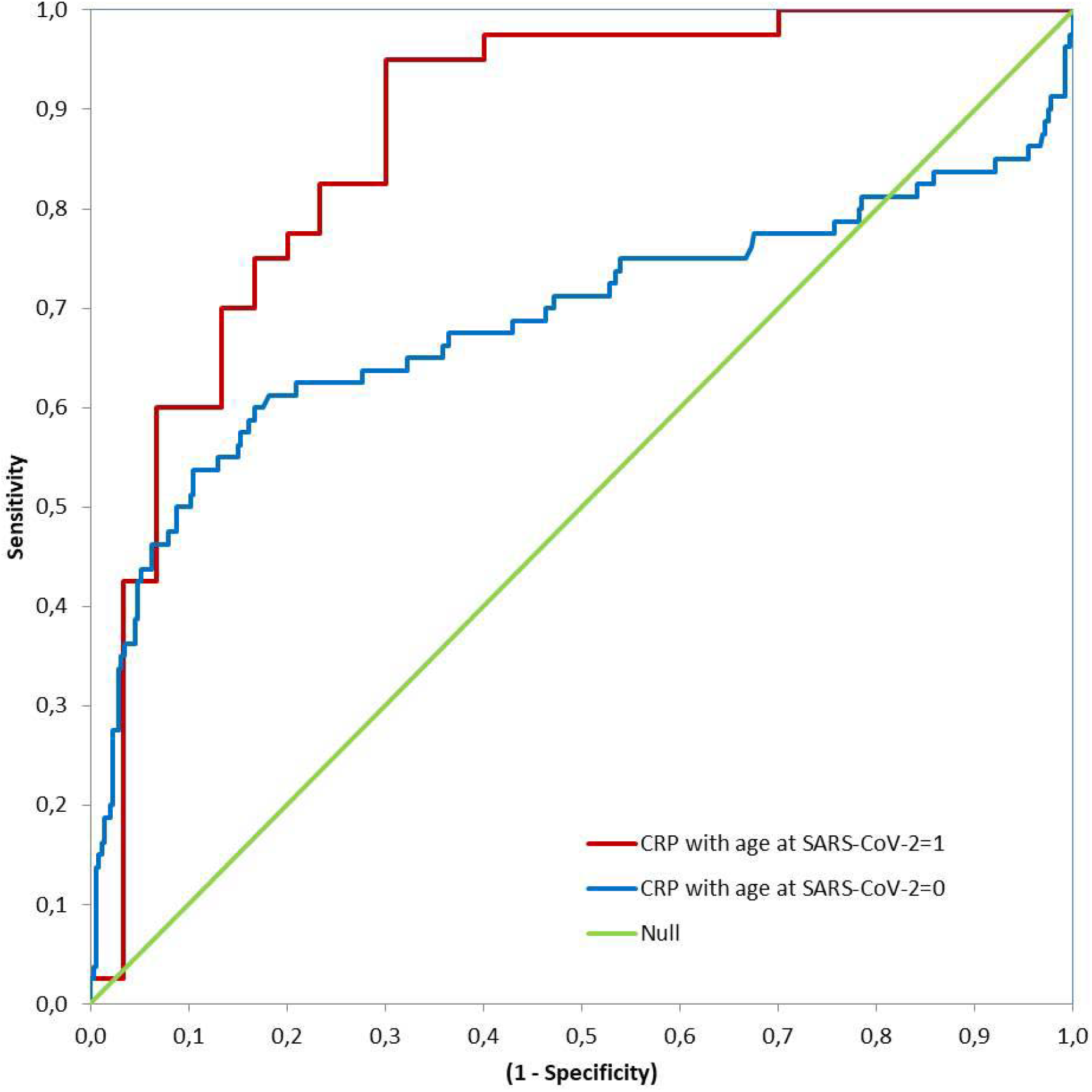
ROC (Receiver Operating Characteristic) curves of the logistic regression model for hospitalization prediction with C-reactive Protein (CRP) controlled for age quantile at both strata (with and without exposure to SARS-CoV-2) Null – Area of the null hypothesis model is 0,5

The Neutrophils to Lymphocytes Ratio (NLR) is considered a possible indicator of severity [21,24,27,28] of COVID-19, but NLR could not be evaluated as all variables were standardized (division by zero). Lymphocytes presented inconsistent behavior across models. Single exam models indicated lymphopenia at SARS-CoV-2=1, as expected [29,30]. But lymphocytes reversed the sign in the model with neutrophils and age (SARS-CoV-2=1), possibly, due to collinearity between them (Pearson correlation of -,925 and -,937 at positive and negative strata, both significant at, 01 (2-tail)). As there are indications of collinearity issues at both strata, lymphocyte and neutrophils should not be in the same model as independent variables, and this is an indication that NLR may be a noisy association with hospitalization. As models with combinations of neutrophils were slightly better than with lymphocyte, lymphocyte was dropped from analysis.

In the second round, combinations of eosinophils, monocytes, and neutrophils with age were tested systematically. Table 2 presents parameters of models combining eosinophils, monocytes, and neutrophils (with age) and the best model with creatinine (as risk factor). Table 3 presents AUCs for each model with the difference of discriminative ability between strata.

**Table 2.**
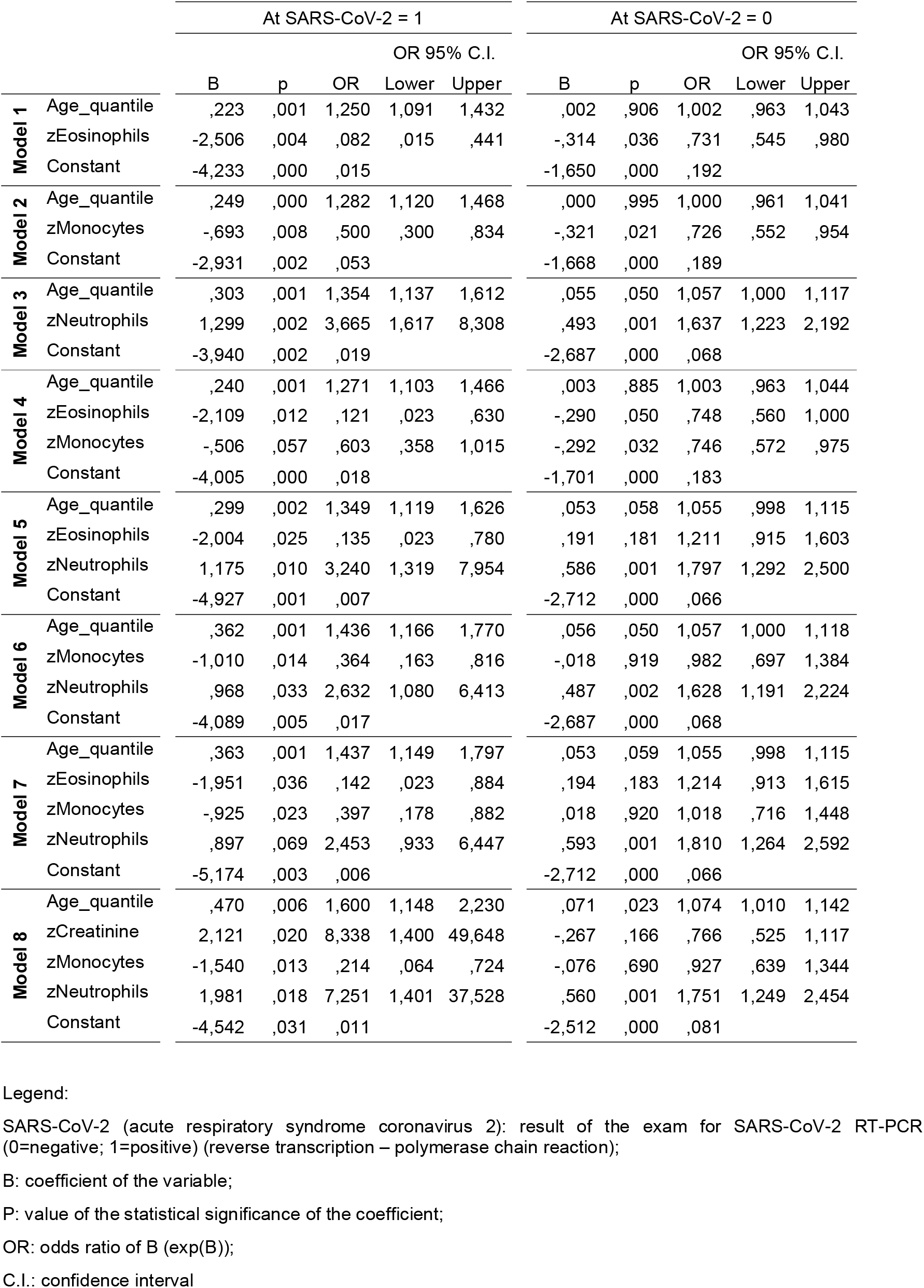
Potential candidate logistic regression models for predicting hospitalization with blood tests and age quantile (different models for each stratum)

**Table 3.** Discriminative ability of potential candidate models for predicting hospitalization from nonspecific blood tests

|  |  | Model |  |  |  |  |  |  |  |
| --- | --- | --- | --- | --- | --- | --- | --- | --- | --- |
|  |  | 1 | 2 | 3 | 4 | 5 | 6 | 7 | 8 |
| Variables included in the model: | zEosinophils | ■ |  |  | • | • |  | • |  |
|  | zMonocytes |  | • |  | • |  | • | • | • |
|  | zNeutrophils |  |  | • |  | • | • | • | • |
|  | Age quantile (0 - 19) | • | • | • | • | • | • | • | • |
|  | Creatinine |  |  |  |  |  |  |  | • |
| Model for positive stratum (SARS-CoV-2=1) | AUC (Area under ROC curve) | ,839 | ,810 | ,862 | ,856 | ,899 | ,897 | ,910 | ,940 |
|  | Standard Error | ,046 | ,049 | ,044 | ,043 | ,036 | ,036 | ,034 | ,029 |
|  | Asymptotic Significance | ,000 | ,000 | ,000 | ,000 | ,000 | ,000 | ,000 | ,000 |
|  | AUC 95% CI |  |  |  |  |  |  |  |  |
|  | Lower bound | ,748 | ,715 | ,775 | ,772 | ,828 | ,826 | ,844 | ,883 |
|  | Upper bound | ,929 | ,906 | ,948 | ,941 | ,970 | ,967 | ,976 | ,997 |
|  | Classification table (cut value = 0,5) |  |  |  |  |  |  |  |  |
|  | Percentage correct H=0 | 70,0 | 70,0 | 75,0 | 75,0 | 72,2 | 72,2 | 75,0 | 81,0 |
|  | Percentage correct H=1 | 79,1 | 79,1 | 84,6 | 83,7 | 87,2 | 82,1 | 89,7 | 82,9 |
|  | Overall percentage | 74,7 | 74,7 | 80,0 | 79,5 | 80,0 | 77,3 | 82,7 | 82,1 |
|  | Cases included in the analysis |  |  |  |  |  |  |  |  |
|  | H=0 | 40 | 40 | 36 | 40 | 36 | 36 | 36 | 21 |
|  | H=1 | 43 | 43 | 39 | 43 | 39 | 39 | 39 | 35 |
|  | Total | 83 | 83 | 75 | 83 | 75 | 75 | 75 | 56 |
| Same model of positive stratum applied to SARS-CoV-2=0 cases | AUC | ,562 | ,542 | ,665 | ,564 | ,603 | ,645 | ,600 | ,627 |
|  | Standard Error | ,036 | ,043 | ,045 | ,037 | ,044 | ,047 | ,044 | ,049 |
|  | Asymptotic Significance | ,069 | ,214 | ,000 | ,061 | ,012 | ,000 | ,016 | ,003 |
|  | AUC 95% CI |  |  |  |  |  |  |  |  |
|  | Lower bound | ,492 | ,459 | ,576 | ,492 | ,518 | ,553 | ,513 | ,532 |
|  | Upper bound | ,632 | ,626 | ,754 | ,636 | ,689 | ,737 | ,686 | ,723 |
|  | Cases included in the analysis |  |  |  |  |  |  |  |  |
|  | H=0 | 433 | 432 | 382 | 432 | 382 | 382 | 382 | 244 |
|  | H=1 | 86 | 86 | 56 | 86 | 56 | 56 | 56 | 55 |
|  | Total | 519 | 518 | 438 | 518 | 438 | 438 | 438 | 299 |
| Difference of the discriminative ability (naïve) |  | 0,277 | 0,268 | 0,197 | 0,292 | 0,295 | 0,252 | 0,310 | 0,313 |
| Overall discriminative performance order |  | 5 | 6 | 8 | 4 | 3 | 7 | 2 | 1 |

AUC: Area under the receiver operating characteristic curve;
CI: confidence interval;
H: Hospitalization (0=false; 1=regular ward, semi-intensive care, or intensive care unit);
SARS-CoV-2 (acute respiratory syndrome coronavirus 2): result of the exam for SARS-CoV-2 RT-PCR (0=negative; 1=positive) (reverse transcription – polymerase chain reaction)

Considered individually, eosinophils, monocytes, and neutrophils generated models with good performance to estimate the probability of hospitalization (models 1, 2, 3 with AUC>,810 at positive stratum). The combinations of these blood tests generated models (4, 5, 6, 7) with better discriminative ability (AUC>,856 at SARS-CoV-2=1). The AUC at SARS-CoV-2=0 is a simplified measure of the systematic bias in both outcomes: models 1, 2, and 4 presented low values (with AUC<,564) and the others presented relevant noisy associations (AUC from, 600 up to, 665), but with better difference in discriminative ability Δ>,252 in models with two or more exams.

Two patterns of associations were more salient: (1) age as a risk factor with combinations of eosinophils, monocytes, and neutrophils as predictors; (2) age and creatinine as risk factors with monocytes and neutrophils as predictors. The interpretation of the conditional probabilities will focus on models 7 and 8, but models with at least two blood tests (4 to 8) are potential candidate associations. Considering creatinine as a marker of the renal function, model 8 is the overall best model with significant coefficients at p<,05 and has the highest difference of discriminative ability between strata (Δ=,313). Comparative ROC curves for models 7 and 8 are shown in Figures 5 and 6, where there is a substantial discriminative difference between both strata of SARS-CoV-2; confidence intervals at 95% of AUC values are in Table 3.

**Figure 5.**
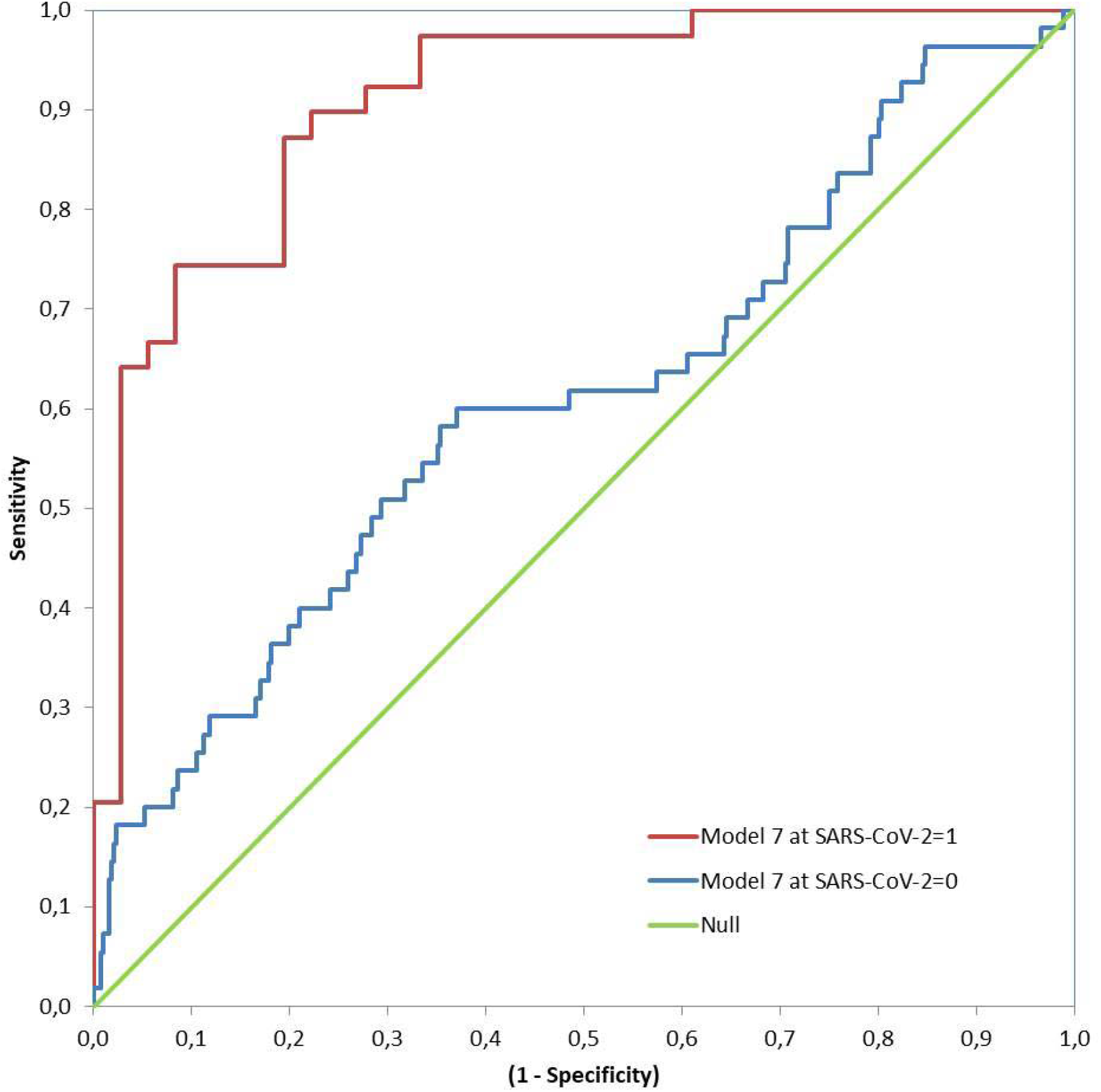
ROC (Receiver Operating Characteristic) curves of model 7 to predict hospitalization at both strata (with and without exposure to SARS-CoV-2) Null – Area of the null hypothesis model is 0,5; Model 7 – Logistic regression with Eosinophils, Monocytes, and Neutrophils controlled for age quantile

**Figure 6.**
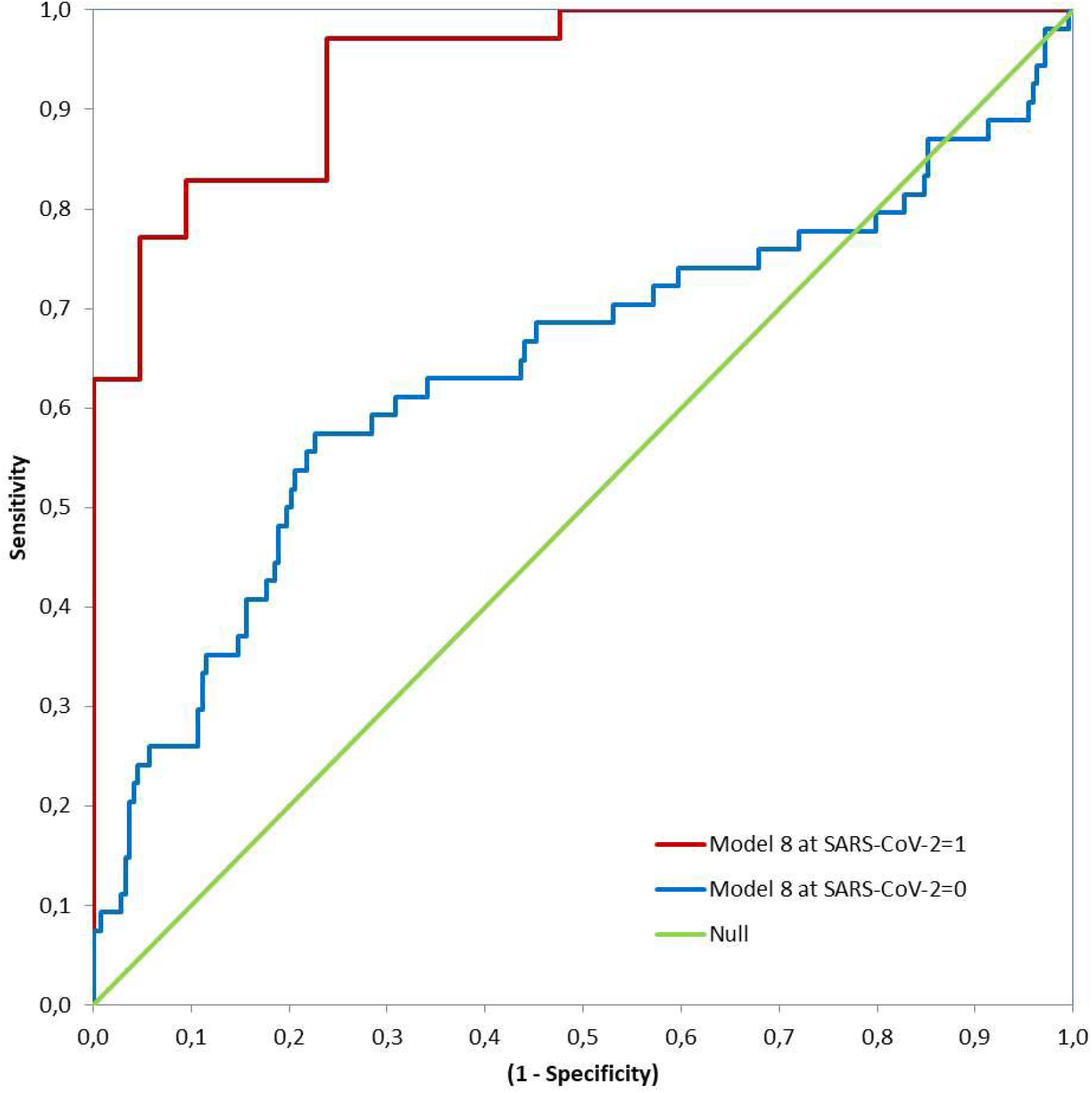
ROC (Receiver Operating Characteristic) curves of model 8 to predict hospitalization at both strata (with and without exposure to SARS-CoV-2) Null – Area of the null hypothesis model is 0,5; Model 8 – Logistic regression with Monocytes and Neutrophils controlled for Creatinine and age quantile

When the coefficients of model 7 (Table 2) are converted to conditional probabilities we find that at average age quantile (9,32) and average monocyte and neutrophil levels, there is a hospitalization probability of 51,1% with eosinophils at -1 standard deviation (SD); and 90,2% when age quantile is 15. Model 8 with creatinine has different responses: age quantile coefficient is more pronounced and the odds ratio of creatinine is steep (8,338), so average levels of creatinine result in a probability of hospitalization >50% for age quantile >9 (with monocytes and neutrophils at average). When creatinine is +1 SD at age quantile 9, hospitalization probability is 85,9% (monocytes and neutrophils at average). Only below average levels of creatinine lower hospitalization probabilities. Monocytes and neutrophils are also steeper than model 7. At age quantile 9, +1/2 SD of creatinine, -1/2 SD of monocytes, and +1/2 SD of neutrophils result in a hospitalization probability of 92,5%.

Model biases may be due to missing cases selection. Most likely, missing data are not at random (MNAR). We performed the bootstrapping procedure to identify potential sensitivity to resampling and, indirectly, to selection bias. The selected models maintained the magnitude and statistical significance of the coefficients. Apparently, as no significant deviation was detected, the missing cases bias may not be an issue.

ROC and AUC calculations used the same data for model fitting. Because of limited sample size, it was not suitable to apply the approach of splitting the database for training and then prediction. After dividing the sample in two groups, most coefficients were not significant at p>0,10 (Table 4) at least in one group. Notwithstanding, classification tables were coherent between sub-sets and we found no clear indication of model misspecification.

**Table 4.**
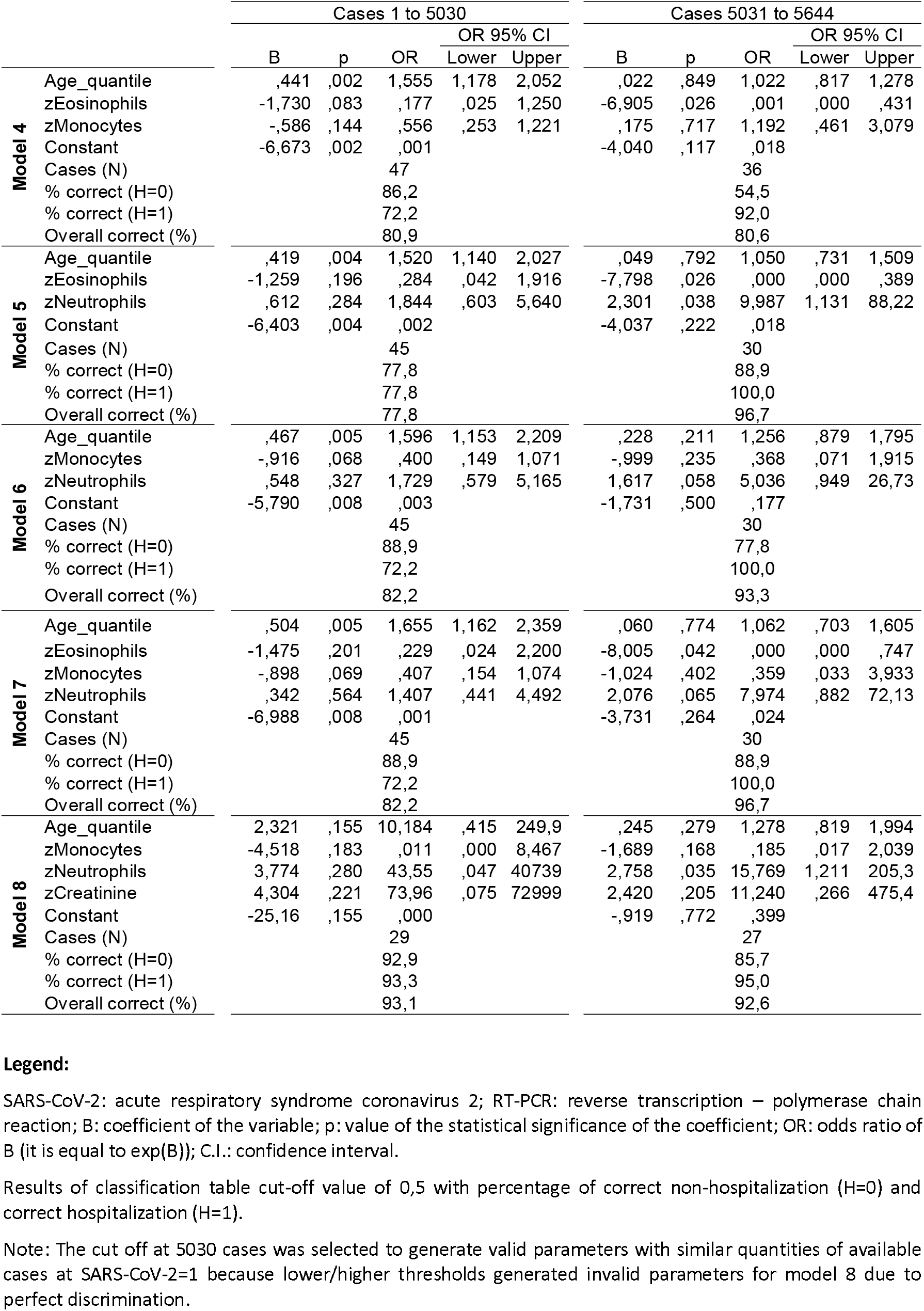
Tentative parameters for models 4 to 8 with split dataset at the positive stratum of SARS-CoV-2: sample size is unsuitable for training and then prediction

## DISCUSSION

We focused on models with discriminative ability to identify peculiar responses in the transition from moderate to severe inflammation only due to COVID-19. The AUC evaluation at the negative SARS-CoV-2 stratum to estimate the influence of unwanted confounders into the focal association together with equivalent criteria of severity state at both strata is, to the best of our knowledge, a needed improvement in prognosis studies of COVID-19.

In comparison to other prediction studies, we identified a few focused on the transition from moderate to severe cases of COVID-19 [21,22,23,24,25,26,27,28]. None of them considered data from the negative stratum of SARS-CoV-2, therefore, these models are biased by not excluding noisy predictors.

We eliminated variables with “high” AUC at SARS-CoV-2=0, so that variables with more peculiar responses to COVID-19 were included. Reactive levels of CRP together with SARS-CoV-2 RT-PCR exam may be a predictor of hospitalization, but this can happen due to causes other than COVID-19 (most cases of COVID-19 are asymptomatic to mild). To include it in a model, one should control for all other causes of CRP reactive.

We evaluated age and creatinine as risk factors. Controlling for age improved the AUC of all models at the positive stratum of SARS-CoV-2. The difference between risk factor and outcome among blood tests is subtle. The emergent literature is cautious about whether eosinopenia may be a risk factor [31] and whether creatinine (and other renal markers) may be associated with COVID-19 renal inflammatory response [32]. As an acute inflammatory kidney response to COVID-19, the interpretation changes and further refinement of the framework is necessary. If eosinopenia is a risk factor, the prevalence of this condition should be considered and must be properly diagnosed at admission, and the models should be reviewed with new data.

As we drop noisy predictors, we are effectively dealing with hypothesis about the physiopathology of COVID-19 inflammation. Although not as frequent as the mentions of neutrophils, there are studies on the complex role of eosinophils [31,33] and monocytes [34,35] in COVID-19 inflammation indicating eosinopenia in severe cases and monocytopenia in some phase of the cytokine storm and other COVID-19 pathologies [36].

We selected two patterns of blood tests that are associated with hospitalization due to COVID-19 inflammation: age with combinations of eosinophils, monocytes and neutrophils; and age and creatinine with monocytes and neutrophils. The model findings are aligned with the known physiopathology of COVID-19 but in a more integrative framework of analysis (not as individual predictors, but as a set that is related to risk factors). The selected blood tests are broadly available even in regions with scarce health care resources. It is unlikely that we will have just one or two overall best models; given different sets of risk factors, we should expect a few representative patterns of the COVID-19 inflammation from moderate to severe.

### Limitations and future directions

The models are candidates only and the results cannot be representative beyond the patient health profiles of this reference hospital in Sao Paulo/Brazil that attends a high social-economic segment [37]. The sample refers to the initial phase of the pandemics in Brazil and the patterns may change with medicine prescriptions and other adaptations of SARS-CoV-2. The reduced quantity of available cases did not allow the dataset split for training and prediction. Further efforts are needed to increase internal and external validity across populations, as the prognostic ability is also a function of the variability of the development of COVID-19 inflammation.

As there is no unambiguous way to characterize “moderate to severe COVID-19 inflammation”, the inclusion of an unmeasured variable reduces the predicted conditional independences from the DAG. But still this framework can help in the identification and estimation of risk factors. This cross-sectional data (single point time) cannot inform if creatinine (or eosinophil) is risk factor or effect of COVID-19 inflammation. In future data collection efforts, participants should be followed over time, from diagnosis to hospitalization; ideally from exposure throughout the lifecycle and also with the follow-up of negative cases.

Causal studies are intrinsically predictive [10], therefore we need to advance prognosis research within causal frameworks. As most studies will be observational, data collection with ample selection of variables for matching estimators (e.g., stratification) [16] will be required to reduce systematic bias.

All candidate models can be reproduced from the dataset [18]. We believe most hospitals can apply this framework to generate similar models appropriate to the target population in which they are inserted by making efforts to collect blood tests and potential risk factors at admission, and other clinical data. By making these databases public (anonymized and with standardized data), they will allow future external validation in larger target populations.

Finally, in the wider context of COVID-19 epidemiology, the collapse of health systems due to opportunistic pathogens is a symptom of threats that requires system-level measures during and after the pandemics [38]. This research is concerned with hospital care. As a bottleneck, even small gains may have multiplicative effects on health systems. In countries with porous containment efforts, hospital occupancy is a critical metric [39] to alternate between “soft lockdown” and economic activity with “constrained mobility”. As some regions with sustained transmission are hesitant and being pushed towards these states, they are poorly capturing the benefits of the switching strategy (Parrondo’s paradox applied to epidemics [40]) – because they are struggling in trial and error mode to establish thresholds of when to restrain (and open) and at what pace. Due to the fast saturation of hospital infra-structures with overshooting in these regions, the tendency of excessive losses in each transition is hard to manage. In this context, we believe that the application of prognosis tools can improve the timely access to supportive care in countries with sustained COVID-19 transmission.

## Data Availability

The data that support the results of this study are openly available.

https://www.kaggle.com/dataset/e626783d4672f182e7870b1bbe75fae66bdfb232289da0a61f08c2ceb01cab01

## Acknowledgements

We are grateful to Antonio Magno Lima Espeschit and Sonia Mara de Andrade who contributed with suggestions to this research. We are also indebted to Hospital Israelita Albert Einstein for making the dataset available, and the referees for their detailed comments.

## Author Contributions

G. Ishikawa: Conceptualization, methodology, and formal analysis

G. Argenti: Conceptualization, formal analysis, and clinical and epidemiological validation

C. B. Fadel: Clinical and epidemiological validation and critical review

All authors: Writing, editing, visualization, review and final approval of manuscript

## Statements

The authors declare no conflicts of interest.

This paper has not been published previously in whole or part.

The data that support the results of this study are openly available in reference number [18].

Although this research received no specific grant from any funding agency, commercial or not-for-profit sectors, as institutionally required we inform that “this study was financed in part by the Coordenacao de Aperfeicoamento de Pessoal de Nivel Superior - Brasil (CAPES) - Finance Code 001”.

